# Right Heart Remodeling After Pulmonary Valve Replacement in Patients with Pulmonary Atresia or Critical Stenosis with Intact Ventricular Septum

**DOI:** 10.1101/2023.05.19.23290256

**Authors:** Margaret Irwin, Lindsey Reynolds, Geoffrey Binney, Stuart Lipsitz, Sunil J Ghelani, David M Harrild, Christopher W Baird, Tal Geva, David W. Brown

## Abstract

**Background:** Patients with pulmonary atresia or critical pulmonary stenosis with intact ventricular septum (PA/IVS) and biventricular circulation may require pulmonary valve replacement (PVR). Right ventricular (RV) remodeling after PVR is well-described in tetralogy of Fallot (TOF); we sought to investigate RV changes in PA/IVS using cardiac magnetic resonance imaging (CMR).

**Methods:** A retrospective cohort of PA/IVS patients who underwent PVR at Boston Children’s Hospital from 1995-2021 with CMR before and after PVR was matched 1:3 with TOF patients by age at PVR. Median regression modeling was performed with post-PVR indexed RV end-diastolic volume (RVEDVi) as the primary outcome.

**Results:** 20 PA/IVS patients (cases) were matched with 60 TOF (controls), with median age at PVR 14 years. Pre-PVR RVEDVi was similar between groups; cases had higher RV ejection fraction (EF; 51.4 vs 48.6%, p=0.03). Pre-PVR RV free wall and LV longitudinal strain (LS) were similar, although LV mid-cavity circumferential strain (CS) was decreased in cases (−15.6 vs -17.1, p=0.001). At median 2 years after PVR, RVEDVi was similarly reduced; cases continued to have higher RV EF (52.3% vs 46.9%, p=0.007) with less reduction in RV mass (Δ4.5 vs 9.6 g/m^2^, p=0.004). Post PVR, RV and LV LS remained unchanged and LV CS was similar, though lower in cases.

**Conclusion:** Compared with TOF patients, PA/IVS patients demonstrate similar RV remodeling after PVR, with lesser reduction in RV mass and comparatively higher RVEF. While no differences were detected in peak systolic RV or LV strain values, further investigation of diastolic parameters is needed.

**Clinical Perspective:**

- This paper provides new insights on the remodeling of the right heart in patients pulmonary atresia with intact ventricular septum or critical pulmonary stenosis, namely that compared with tetralogy of Fallot patients, these patients demonstrate overall similar right ventricular remodeling following pulmonary valve replacement.
- CMR strain imaging found no significant pre-post pulmonary valve replacement differences in right ventricular or left ventricular systolic parameters

## Introduction

Patients with pulmonary atresia or critical pulmonary stenosis (CPS) with intact ventricular septum (PA/IVS) typically have right ventricular hypertrophy, hypoplasia, and both systolic and diastolic dysfunction at birth. The preferred treatment is the establishment of antegrade pulmonary blood flow through interventional catheterization or surgical techniques, which creates the potential for progressive right ventricular (RV) dilation in the absence of a competent pulmonary valve and severe pulmonary regurgitation (PR)^1-7^. Chronic volume load on the RV results in dilation and eventual dysfunction of the ventricle and pulmonary valve replacement (PVR) is required in majority of these patients^8, 9^.

Changes in RV size and function following PVR have been well studied in patients with tetralogy of Fallot (TOF)^10-13^, however, similar data are not available for pateints with PA/IVS undergoing PVR. The goal of PVR in patients with both PA/IVS and TOF is to preserve myocardial function and reverse the RV dilation resulting from the volume load resulting from chronic PR. Understanding the remodeling response to PVR in this population is important as the timing of PVR is often empiric, imaging based, and commonly performed in asymptomatic patients. In absence of data specific to PA/IVS, decisions regarding timing of PVR often rely on studies on patients with TOF. Hence this study seeks to characterize volumetric and functional changes in the RV after PVR in patients with PA/IVS compared to those with TOF.

## Methods

### Patients

We conducted a single-center retrospective cohort review of patients with PA/IVS or CPS who underwent PVR for severe PR at Boston Children’s Hospital (BCH) from 1995-2021. Native pulmonary valve anatomy and ventricular septum status was determined by initial preoperative echocardiography. CPS was defined as ductal-dependent circulation and/or initial intervention on the pulmonary valve at age <2 weeks. Patients managed with single ventricle circulation or a subsequent 1.5 ventricle repair were excluded; those without CMR imaging <5 years prior to and following PVR were also excluded. PA/IVS patients were then matched in a 1:3 ratio with control TOF patients by age (± 1 year) at PVR, with similar requirement for pre- and post-PVR CMR imaging at our institution. The study was approved by the Institutional Review Board at BCH, and individual patient consent was waived. The data that support the findings of this study are available from the corresponding author upon request.

### Demographic, Surgical, and Electrophysiological Data

Baseline and clinical characteristics were extracted from the medical record including demographic information, presence of a genetic disorder, weight, and body surface area at the time of PVR. The following anatomic variables were recorded: native pulmonary valve morphology, status of the ventricular septum, and additional anatomic anomalies. Anatomic criteria for PA/IVS and CPS patients were confirmed in both the pre-operative echocardiogram report and operative note. The following pre-operative surgical and electrophysiological data were recorded for PA/IVS cohort and TOF controls: age at initial pulmonary repair, age at and type of prior cardiac interventions, time between initial pulmonary repair and PVR, and presence of arrhythmia on Holter monitor. Additionally, operative notes were reviewed to collect data about PVR implantation, including surgical techniques, age at PVR, valve size, valve morphology, presence of tricuspid valve repair, additional interventions, and status of the atrial septum. Post-operative data included documented cardiac reintervention, age, clinical status, and New York Heart Status evaluation at most recent follow-up.

### Echocardiography

Echocardiographic data were obtained from clinical reports, or, if missing, on secondary review of primary images (DWB). Data collected included qualitative assessments of RV and left ventricular (LV) function and degree of dilation, pulmonary and tricuspid valve regurgitation (TR), and quantitative assessments of RV systolic pressure and pulmonary stenosis. Qualitative ventricular dysfunction or dilation was characterized as none or trivial, mild, moderate, or severe.

### Cardiovascular Magnetic Resonance

CMR imaging data were collected from clinical reports, or, if missing, on secondary review of images (SG); data included LV and RV ejection fractions, end-diastolic and systolic volumes, mass, and pulmonary and TR fractions. Volumetric data were indexed to body surface area.

Details of the CMR protocol have been previously published.^14^ Briefly, all CMRs were performed on a 1.5 T scanner (Achieva, Phillips Healthcare, Best, the Netherlands) using surface coils selected based on patient size. Imaging included a 12–14 slice stack (slice thickness 8–10 mm) of breath-hold, ECG-gated, balanced steady-state free precession (bSSFP) cine acquisitions in the short-axis plane to completely cover both ventricles. Twenty images per cardiac cycle were acquired and reconstructed to 30 images per cardiac cycle. Feature tracking analysis was performed using commercial software (cvi42, Circle Cardiovascular Imaging v5.13 Calgary, AB, Canada). The following strain variables were collected: RV longitudinal strain (free wall strain on a 4-chamber view); LV longitudinal strain (4-chamber view); and LV circumferential strain (mid-cavity). In order to perform the analysis, ventricular endocardial and epicardial contours were manually drawn at one phase of the cardiac cycle and automatically propagated for the full cardiac cycle by the software. Tracking was manually inspected and the contours were manually adjusted in case of poor tracking.

### Statistical Analysis

Descriptive statistics were used to summarize variables. Frequencies and percentages for categorical variables, and mean ± standard deviation and median (interquartile range) for continuous and ordinal variables were provided as appropriate. Categorical patient characteristics were compared between the PA/IVS and TOF groups using a Rao-Scott Chi-squared test,^15^ and continuous patient characteristics were compared using a Mann-Whitney-Wilcoxon test,^16^ accounting for clustering due to matching respectively.

Because the main outcome data were not distributed normally, generalized estimating equations for the median^17, 18^ (taking into account clustering due to matching) were fit with a fixed effect for diagnosis (PA/IVS versus TOF), and with reduction (change) in indexed right ventricular end diastolic volume (RVEDVi) post PVR as the primary outcome. Secondary analyses provided covariate-adjusted estimates of the diagnosis effect by fitting the model above adjusting for 2 covariates deemed as potential confounding variables: PVR size and degree of TR. These analyses were repeated for the secondary outcomes.

In order to have 80% power (with a 5% Type 1 error) to detect a ratio of medians of 1.4 in the 2 groups, a 1:3 PA/IVS subject to TOF match design was selected for this study. In this power calculation, the test statistic used is a robust generalized estimating equations Wilcoxon rank-sum test (not assuming normality and accounting for clustering due to matching), and we assume an intracluster (matching) correlation coefficient of 0.01 and a coefficient of variation of 0.6. We considered a two-sided p-value ≤0.05 as statistically significant. All statistical analyses were performed using SAS version 9.4 (SAS Institute, Cary, NC).

## Results

### Patient Characteristics

A total of 20 PA/IVS or CPS patients met study inclusion criteria. Baseline patient characteristics are listed in Table 1. PA/IVS was the primary diagnosis in 15 subjects, while 5 had CPS; median age at PVR was 14.8 years [5, 32] and 12 patients were male (60%). Initial neonatal intervention was surgical in 15 (75%), consisting of Blalock-Taussig-Thomas (BTT) shunt in 5, BTT shunt with RV outflow tract patch in 3, and both in 8. Initial neonatal intervention was transcatheter in 9 patients (45%), with pulmonary valve perforation and balloon dilation. Of the patients managed with initial catheter intervention, 4 underwent additional surgery in the first month of life, with RV outflow tract patch in 4 as well as BTT shunt in 2 of these. For the entire cohort, 6 (30%) had an additional surgical interventions following initial repair, including TV plasty in 3 (15%).

**Table 1.**
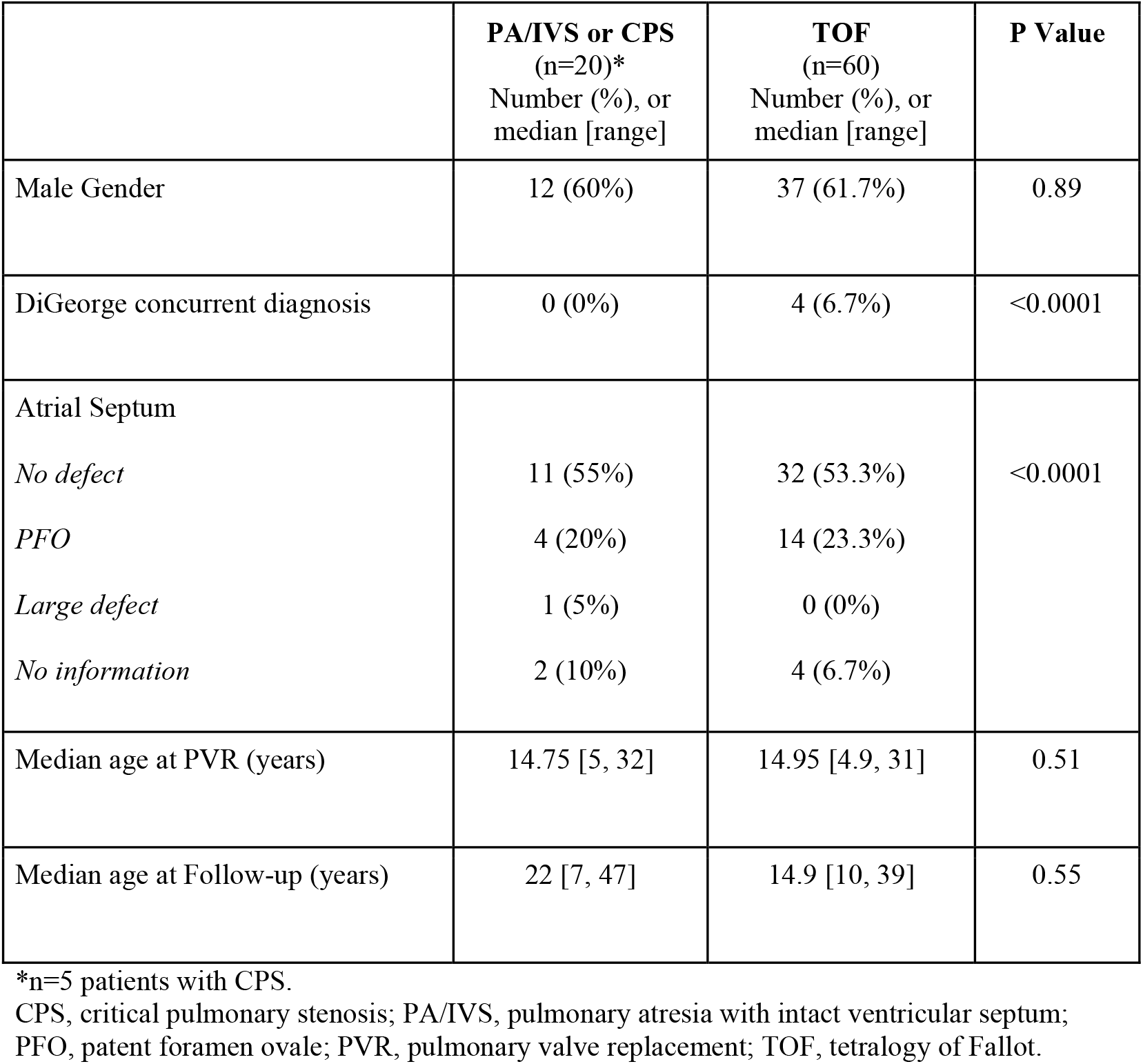
Patient Characteristics.

|  | **PA/IVS or CPS**<br>(n=20)*<br>Number (%), or<br>median [range] | **TOF**<br>(n=60)<br>Number (%), or<br>median [range] | **P Value** |
| --- | --- | --- | --- |
| Male Gender | 12 (60%) | 37 (61.7%) | 0.89 |
| DiGeorge concurrent diagnosis | 0 (0%) | 4 (6.7%) | <0.0001 |
| Atrial Septum |  |  |  |
| *No defect* | 11 (55%) | 32 (53.3%) | <0.0001 |
| *PFO* | 4 (20%) | 14 (23.3%) |  |
| *Large defect* | 1 (5%) | 0 (0%) |  |
| *No information* | 2 (10%) | 4 (6.7%) |  |
| Median age at PVR (years) | 14.75 [5, 32] | 14.95 [4.9, 31] | 0.51 |
| Median age at Follow-up (years) | 22 [7, 47] | 14.9 [10, 39] | 0.55 |

\*n=5 patients with CPS.
CPS, critical pulmonary stenosis; PA/IVS, pulmonary atresia with intact ventricular septum; PFO, patent foramen ovale; PVR, pulmonary valve replacement; TOF, tetralogy of Fallot.

A total of 60 TOF patients who met inclusion criteria were matched with PA/IVS patients. Pre-PVR interventions for this cohort included TOF repair at median 0.2 years, with transannular patch repair in 43 (75.4%); 5 patients underwent BTT shunt procedure prior to TOF repair. Additional surgeries following TOF repair for this group prior to PVR were performed in 19 of 60 patients (32%), including RVOT patch revision in 7 (11%), and RV-PA conduit revision in 6 (10%).

### Pre-PVR Imaging

Pre-operative echocardiographic data are summarized in Table 2. There were no significant differences between PA/IVS and TOF patients with regard to frequencies of ≥ moderate RV dilation, ≥ moderate RV dysfunction, ≥ moderate LV dysfunction, and ≥ moderate pulmonary regurgitation (PR). Moderate or severe TR was more common in PA/IVS patients (80%) as compared with those with TOF (5%) (p =0.01).

**Table 2.** Echocardiographic Data.

|  | **PA/IVS or CPS\***<br>(n=20)<br>Number (%) | **TOF**<br>(n=60)<br>Number (%) | **P Value** |
| --- | --- | --- | --- |
| **Pre-PVR** |  |  |  |
| RV dilation ≥ moderate | 16 (80) | 44 (73) | 0.61 |
| RV dysfunction ≥ moderate | 0 (0) | 3 (5) | 0.94 |
| LV dysfunction ≥ moderate | 0 (0) | 0 (0) | - |
| PR ≥ moderate | 20 (100) | 50 (83) | 0.93 |
| TR ≥ moderate | 16 (80) | 3 (5) | 0.001 |
| **Post- PVR** |  |  |  |
| RV dilation ≥ moderate | 4 (20) | 12 (20) | 0.98 |
| RV dysfunction ≥ moderate | 0 (0) | 4 (7) | 0.93 |
| LV dysfunction ≥ moderate | 0 (0) | 2 (3) | 0.95 |
| PR ≥ moderate | 6 (30) | 6 (10) | 0.16 |
| TR ≥ moderate | 7 (35) | 3 (5) | 0.014 |

\*n=5 patients with CPS.
CPS, critical pulmonary stenosis; LV, left ventricular; PA/IVS, pulmonary atresia with intact ventricular septum; PFO, patent foramen ovale; PR, pulmonary regurgitation; PVR, pulmonary valve replacement; RV, right ventricular; TOF, tetralogy of Fallot; TR, tricuspid regurgitation.

Pre-PVR CMR data and CMR strain values are summarized in Tables 3 and 4. Pre-PVR RVEDVi was similar between the PA/IVS and TOF cohorts (165 vs. 167 ml/m^2^, p=0.7). PA/IVS patients had higher RV EF (51.4 vs. 48.6%, p=0.014), TR fraction (23 vs. 9%, p=0.008), and a trend toward lower indexed RV mass (32.5 vs. 35.6 g/m2, p=0.08). Notably, PR fraction, LV volumes, and LV EF were similar. Pre-PVR strain data demonstrated similar peak RV longitudinal strain (p=0.9) and LV longitudinal strain (p=0.5) between groups, although LV mid-cavity circumferential strain was reduced in the PA/IVS group compared with TOF controls (−15.6 vs. -17.1%, p=0.0008).

**Table 3.**
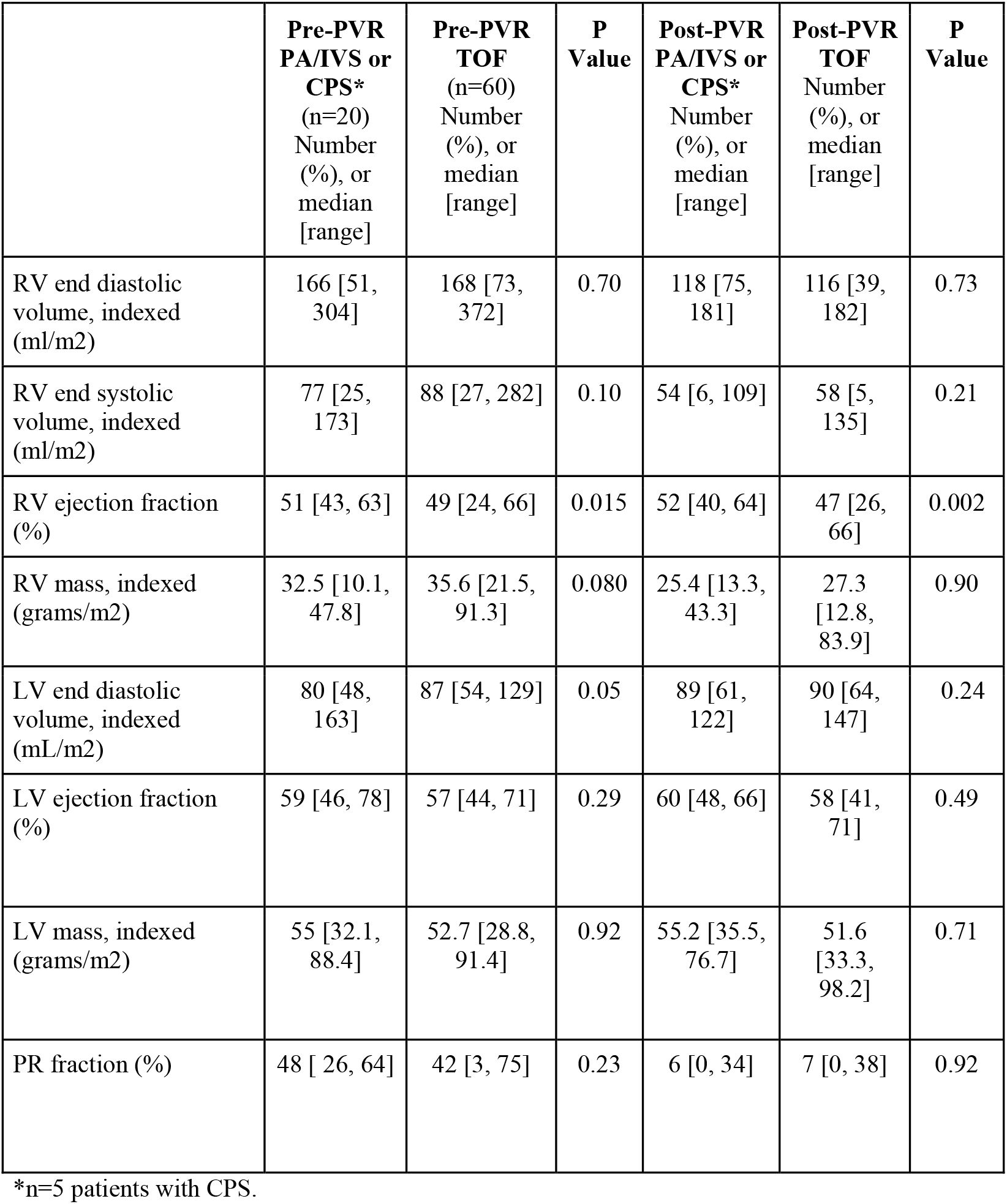
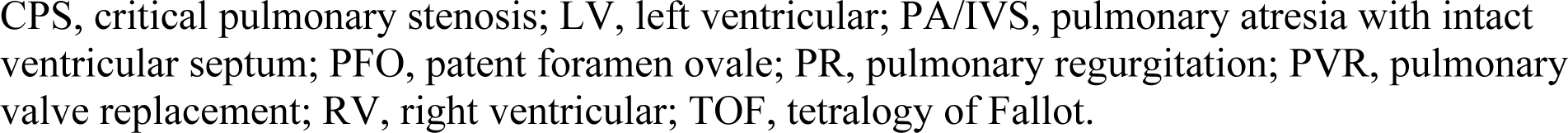
Cardiac Magnetic Resonance Volumetric and Function Data.

|  | **Pre-PVR**<br>**PA/IVS or**<br>**CPS\***<br>**(n=20)**<br>**Number**<br>**(%), or**<br>**median**<br>**[range]** | **Pre-PVR**<br>**TOF**<br>**(n=60)**<br>**Number**<br>**(%), or**<br>**median**<br>**[range]** | **P**<br>**Value** | **Post-PVR**<br>**PA/IVS or**<br>**CPS\***<br>**Number**<br>**(%), or**<br>**median**<br>**[range]** | **Post-PVR**<br>**TOF**<br>**Number**<br>**(%), or**<br>**median**<br>**[range]** | **P**<br>**Value** |
| --- | --- | --- | --- | --- | --- | --- |
| RV end diastolic volume, indexed (ml/m<sup>2</sup>) | 166 [51, 304] | 168 [73, 372] | 0.70 | 118 [75, 181] | 116 [39, 182] | 0.73 |
| RV end systolic volume, indexed (ml/m<sup>2</sup>) | 77 [25, 173] | 88 [27, 282] | 0.10 | 54 [6, 109] | 58 [5, 135] | 0.21 |
| RV ejection fraction (%) | 51 [43, 63] | 49 [24, 66] | 0.015 | 52 [40, 64] | 47 [26, 66] | 0.002 |
| RV mass, indexed (grams/m<sup>2</sup>) | 32.5 [10.1, 47.8] | 35.6 [21.5, 91.3] | 0.080 | 25.4 [13.3, 43.3] | 27.3 [12.8, 83.9] | 0.90 |
| LV end diastolic volume, indexed (mL/m<sup>2</sup>) | 80 [48, 163] | 87 [54, 129] | 0.05 | 89 [61, 122] | 90 [64, 147] | 0.24 |
| LV ejection fraction (%) | 59 [46, 78] | 57 [44, 71] | 0.29 | 60 [48, 66] | 58 [41, 71] | 0.49 |
| LV mass, indexed (grams/m<sup>2</sup>) | 55 [32.1, 88.4] | 52.7 [28.8, 91.4] | 0.92 | 55.2 [35.5, 76.7] | 51.6 [33.3, 98.2] | 0.71 |
| PR fraction (%) | 48 [ 26, 64] | 42 [3, 75] | 0.23 | 6 [0, 34] | 7 [0, 38] | 0.92 |

\*n=5 patients with CPS.
CPS, critical pulmonary stenosis; LV, left ventricular; PA/IVS, pulmonary atresia with intact ventricular septum; PFO, patent foramen ovale; PR, pulmonary regurgitation; PVR, pulmonary valve replacement; RV, right ventricular; TOF, tetralogy of Fallot.

**Table 4.** Cardiac Magnetic Resonance Strain Data.

|  | **Pre-PVR**<br>**PA/IVS or**<br>**CPS\***<br>**(n=20)**<br>**median**<br>**[range]** | **Pre-PVR**<br>**TOF**<br>**(n=60)**<br>**median**<br>**[range]** | **P**<br>**Value** | **Post-**<br>**PVR**<br>**PA/IVS**<br>**or**<br>**CPS\***<br>**(n=20)**<br>**median**<br>**[range]** | **Post-PVR**<br>**TOF**<br>**(n=60)**<br>**median**<br>**[range]** | **P**<br>**Value** |
| --- | --- | --- | --- | --- | --- | --- |
| RV longitudinal strain | -18.8 [-10.9, -24.8] | -18.8 [-7.6, -25.8] | 0.88 | -18.1 [-10.9, -22.4] | -18.1 [-7.3, -26.0] | 0.21 |
| LV mid-cavity circumferential strain | -15.6 [-9, -18.8] | -17.1 [-12.1, -22.4] | 0.001 | -15.9 [-12.3, -20.4] | -18.7 [-12.1, -22.6] | 0.001 |
| LV longitudinal strain | -14.9 [-9.7, -22.4] | -14.8 [-6.9, -21.7] | 0.50 | -14.7 [-11.9, -19.1] | -15.7 [-7.3, -22.0] | 0.18 |

\*n=5 patients with CPS.
CPS, critical pulmonary stenosis; LV, left ventricular; PA/IVS, pulmonary atresia with intact ventricular septum; PVR, pulmonary valve replacement; RV, right ventricular; TOF, tetralogy of Fallot.

### PVR Data

Median age at PVR was similar between groups, 14.7 years for the PA/IVS and 14.9 years for the TOF group (p=0.51). Tricuspid valve repair was performed with PVR surgery in 14 PA/IVS subjects (70%) and 7 TOF patients (12%) (p=0.001). All patients received a bioprosthetic pulmonary valve, with the most common being Carpentier Edwards Magna in 33 (41%) and Sorin Mitroflow in 26 (33%). The median valve size was similar between PA/IVS (25 mm) and TOF (24 mm) cohorts (p=0.12). In terms of placement technique, one patient (5%) in the PA/IVS cohort had transcatheter PVR, and 11 patients (18%) in the TOF cohort had transcatheter PVR (p<0.001).

### Post PVR Imaging Data

Post-PVR echocardiographic imaging was performed at median 18.6 months post PVR with a statistically significant difference between groups in timing (33 months for PA/IVS vs. 12 months for TOF, p=0.004) (Table 2). Findings included similar reduction in RV size, with ≥ moderate dilation present in 20% of both PA/IVS and TOF cohorts following PVR (p=0.98). No PA/IVS subjects had ≥ moderate post-PVR RV dysfunction by echocardiography, compared to 4 TOF patients (7%, p=0.93); similarly, no PA/IVS subjects had ≥ moderate LV dysfunction post PVR, compared with 2 TOF patients (3%, p=0.95). However, TR was more common in the PA/IVS cohort, with ≥ moderate TR in 35% versus only 5% of TOF patients (p=0.014).

CMR data were obtained at a median of 2 years after PVR, with no significant difference in timing between cohorts (2.85 years for PA/IVS vs 1.65 years for TOF cohort, p=0.13). Both cohorts had significant and similar reductions in RVEDVi after PVR, from 166 to 118 ml/m^2^ in the PA/IVS group and from 168 ml/m^2^ to 116 ml/m^2^ in the TOF group (Table 3). PA/IVS patients had higher post-PVR RV EF (52.3% vs. 46.9%, p=0.007) with less reduction in indexed RV mass (Δ4.5 vs 9.6 g/m^2^, p=0.004) compared to the TOF group. LV volumes and LV EF were similar.

CMR strain analyses post-PVR (Table 4) demonstrated that RV free wall longitudinal strain was similar between PA/IVS and TOF patient groups after PVR (−18.1 vs -18.1, p=0.2). Moreover, LV longitudinal strain also remained similar both within and between PA/IVS versus TOF groups (−14.7 vs -15.7, p 0.18). However, the LV mid-cavity circumferential strain remained reduced in PA/IVS versus TOF patients post-PVR (−15.9 vs -18.7, p=0.001), with no significant pre-post PVR differences within groups (Δ -0.3 vs -1.6, p=0.47).

### Post PVR Follow-up Clinical Data

Median age at last clinical follow-up was 22 years for both cohorts, with no statistically signficant difference in follow-up time between cohorts (p=0.55). There was no mortality after PVR in either PA/IVS or TOF cohorts. At follow-up, 95% of both PA/IVS or CPS and TOF patients were in New York Heart Association status Class 1 (p=0.99). No patients in either group were diagnosed with atrial or ventricular tachyarrhythmias, including by surveillance 24-hour Holter monitor testing performed in 60% of PA/IVS patients and 45% of TOF patients. Exercise stress tests were performed in a subset of both cohorts; paired pre/post-PVR exercise tests were available in 10 PA/IVS subjects and 18 age-matched TOF control patients with similarly paired testing. There were no statistically significant between group differences in pre- and post-PVR exercise parameters (peak and % predicted peak V0_2_, heart rate, and 02 pulse).

## Discussion

In this study, we sought to characterize volumetric and functional changes in the RV after PVR in patients with PA/IVS and CPS using CMR imaging with comparison to a control group of age-matched TOF patients. In this cohort of 20 patients, we found that volumetric changes following PVR were remarkably similar to TOF controls, with similar marked reductions in RVEDVi. RV indexed mass changed less in PA/IVS patients after PVR than in TOF controls, although TOF controls had generally greater pre-PVR RV mass to begin with. Similarly, RV EF was slightly better in PA/IVS patients than TOF controls post-PVR, although that was also true pre-PVR as well. LV volumes and EF did not significantly change in either group after PVR.

A number of large cohort studies in the TOF population have demonstrated the impact of PVR on the RV, with generally consistent findings that the RV will often remodel with 30-40% reduction in RVEDV, and generally no improvement in RV or LV EF.^10^ Prior studies have shown that RV response to volume overload is different for patients with PA/IVS^8^. One single-center study compared the effect of PVR in PA/IVS patients with TOF patients^19^; however, this study was limited by lack of post-PVR cardiac magnetic resonance (CMR) imaging follow up (only 5/13 patients) and focused largely on tricuspid regurgitation (TR) and RV size. Our study confirms that the same volumetric changes occur after PVR in the PA/IVS population to those with TOF, albeit with slightly better pre- and post-PVR RV EF. Importantly, these similar volumentric changes in the PA/IVS group occurred in the setting of significant TR in a majority of patients, and this residual TR may be masking what would otherwise be more significant reductions in RV volume after PVR. Further analysis with a larger cohort of subjects (such as a multicenter study) may yield additional insight into this issue.

The present study also demonstrated that PA/IVS patients had no significant change in any of the strain components (including LV and RV longitudinal, or LV circumferential) following PVR. The lack of change in RV strain parameters in PA/IVS is consistent with prior published data in TOF patients^10^, demonstrating again similarities in RV functional characteristics in these 2 populations in response to PVR. However, TOF patients in published reports have been demonstrated to have some improvement in LV strain parameters following PVR^10^, which we did not find in the present study in either group. This is perhaps related to the small sample sizes in our study; the finding of reduced circumferential strain in PA/IVS patients is an interesting observation that deserves further investigation.

### Limitations

Due to the retrospective design, our study has a number of limitations, perhaps most importantly the enrollment criteria for both subject and controls of CMR data within specified time frames pre and post PVR. This bias may be mitigated by the fact that CMR has become a frequently used diagnostic tool for the clinical management of patients with TOF, and by extension, for PA/IVS patients with severe PR and dilated RVs. The small number of patients with CPS included in the study cohort is technically a different cardiac diagnosis than membranous PA/IVS; however, these diagnoses are so closely related in cardiac physiology, neonatal management and subsequent need for potential PVR for severe PR and dilated RV that they were included in the present study. Additionally, a small number of CMR images were not of sufficient quality for strain analysis. Finally, the CMR RV strain measured in this study was the 4-chamber free-wall longitudinal strain, which represents only a portion of the right ventricle and thus may not be able to detect other changes that may have occurred post PVR.

## Conclusions

Patients with PA/IVS with dilated right ventricles demonstrate similar RV remodeling post PVR to TOF patients, with similar reduction in RVEDV, albeit lesser reduction in RV mass and slightly better post PVR RV EF. CMR strain imaging found no significant within or between group differences pre-post PVR in RV or LV systolic parameters. These data offer support to the common clinical practice of basing the timing of PVR in patients with PA/IVS on studies from patients with repaired TOF. However, further investigation is needed to evaluate other metrics such as differences in diastolic function that may influence long-term outcomes.

**Figure 1AB.**
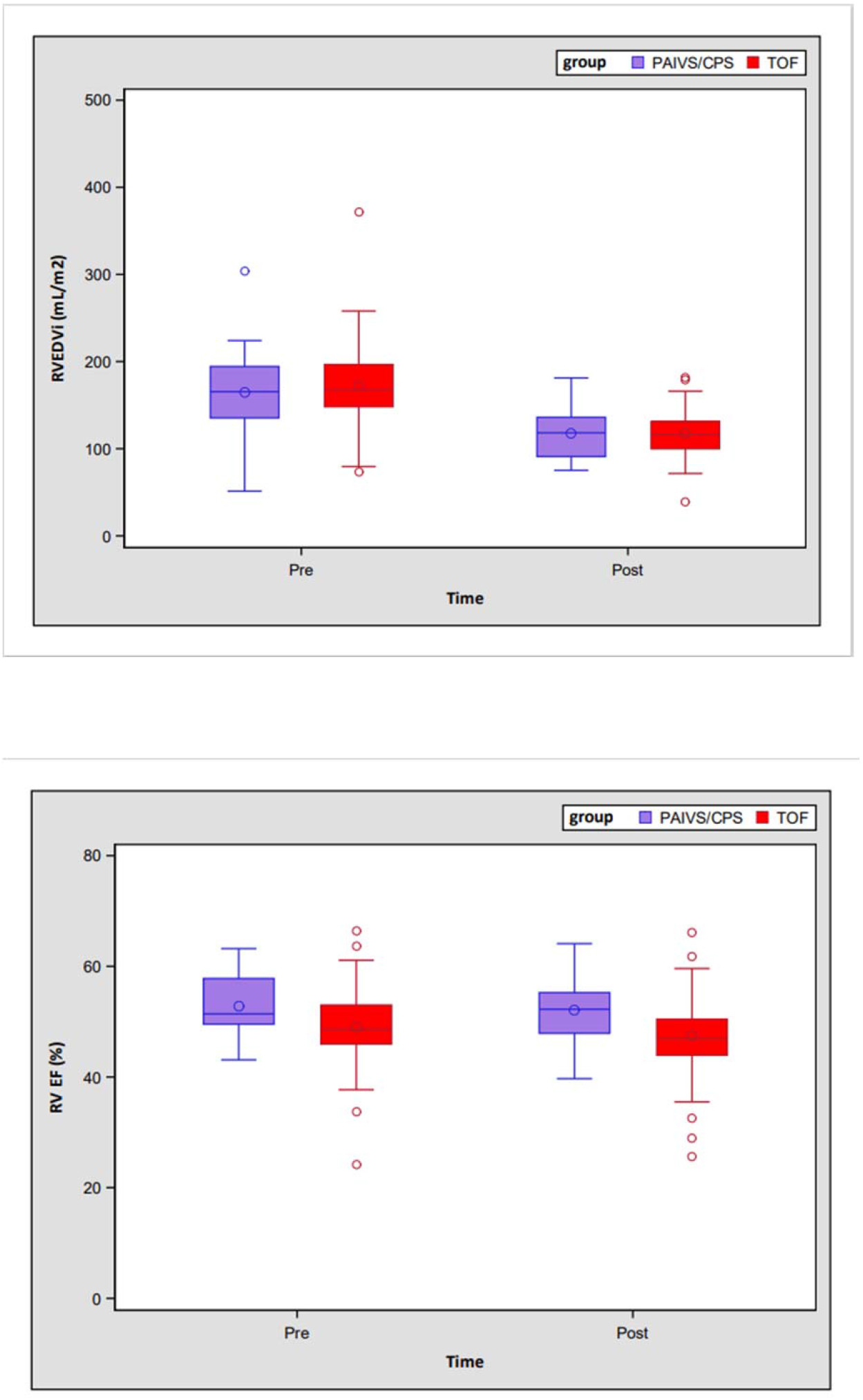
Volumetric Comparison Between Pulmonary Atresia With Intact Ventricular Septum and Tetralogy of Fallot by Cardiac Magnetic Resonance. Box and whiskers plots depict the A) right ventricular end-diastolic volume indexed for body surface area (RVEDVi, y axis), and B) right ventricular ejection fraction (RVEF, y axis) prior to and following pulmonary valve replacement (PVR) for patients with pulmonary atresia with intact ventricular septum (PAIVS, purple) and tetralogy of Fallot (TOF, red). Boxes represent medians with whiskers for IQR.

**Figure 2AB.**
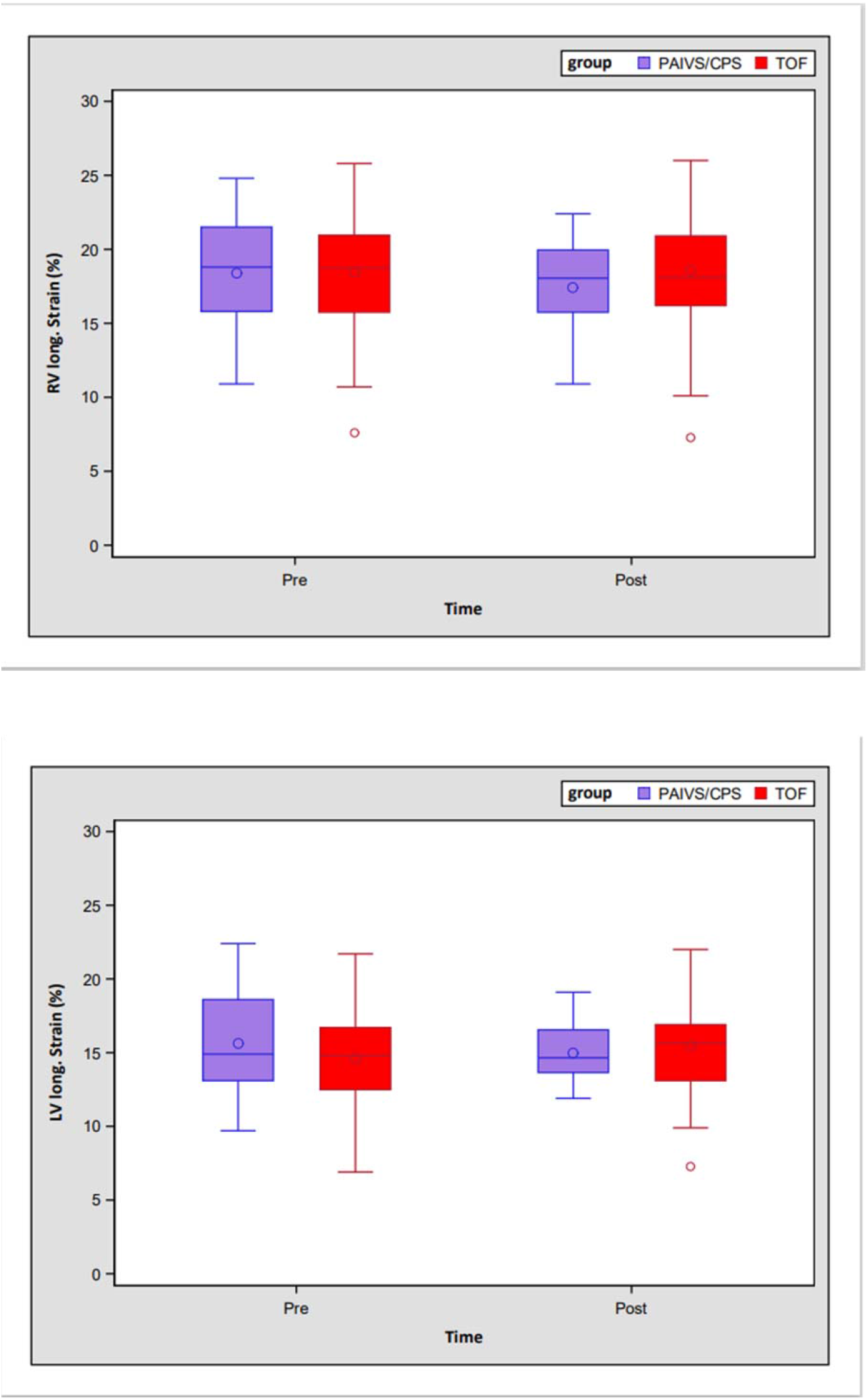
Ventricular Strain Parameters by Cardiac Magnetic Resonance Between Pulmonary Atresia With Intact Ventricular Septum and Tetralogy of Fallot. Box and whiskers plots depict the A) right ventricular (RV) longitudinal strain, and B) left ventricular (LV) longitudinal strain prior to and following pulmonary valve replacement (PVR) for patients with pulmonary atresia with intact ventricular septum (PAIVS, purple) and tetralogy of Fallot (TOF, red). Box is median with whiskers for IQR.

**Figure 3A-C.**
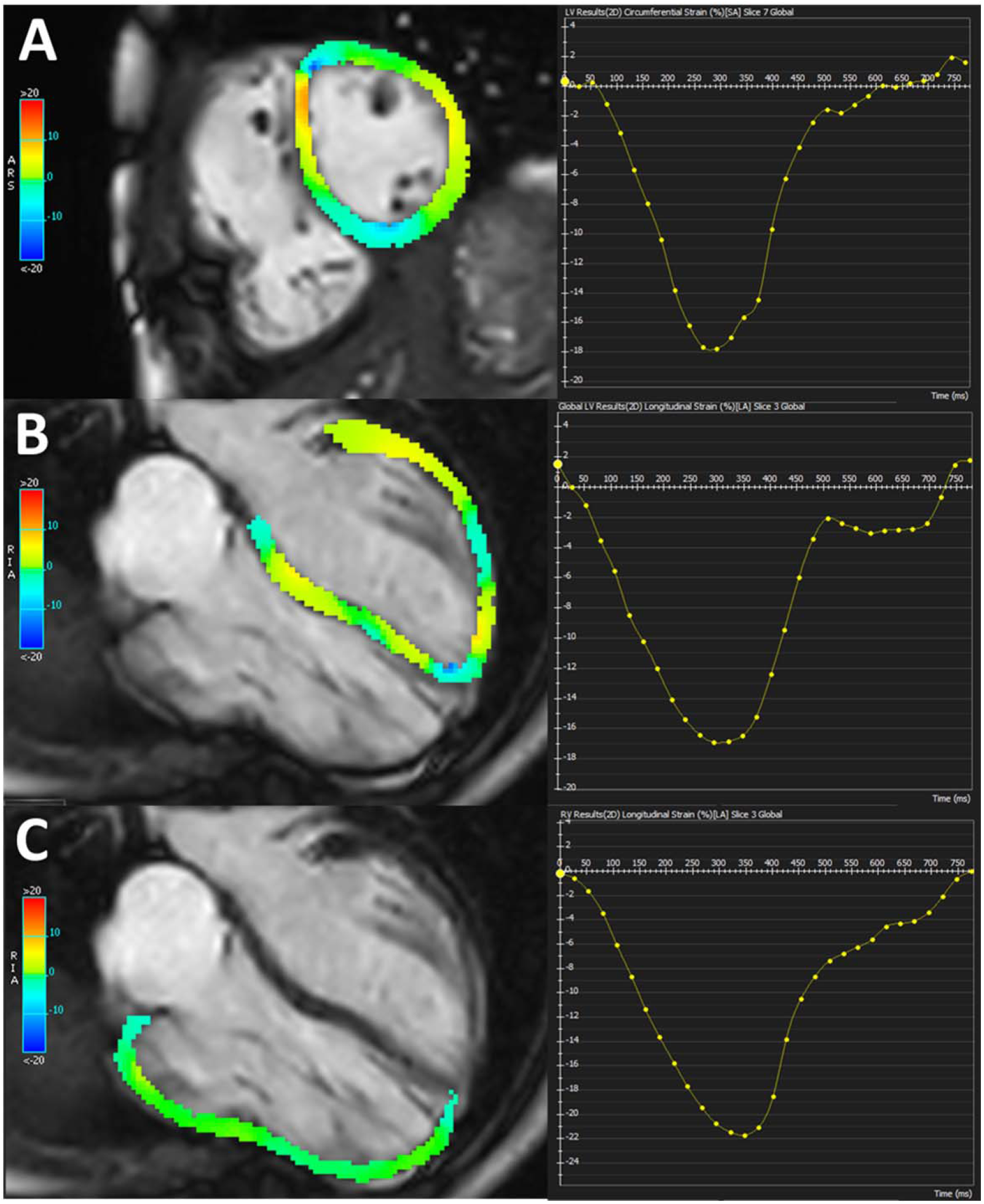
Cardiac Magnetic Imaging Strain Measurement Technique. Cardiac magnetic resonance images in A) 2-chamber short axis, and B,C) 4-chamber long axis views depict the border detection technique used in deriving left ventricular (LV) and right ventricular strain. Longitudinal strain is derived from 4-chamber views, and LV circumferential strain from short axis views. Right-hand panels depict the average strain (negative deflection) of the summated segments over one cardiac cycle.

## Data Availability

The data that support the findings of this study are available from the corresponding author upon request.

## Abbreviations

BTT: Blalock-Taussig-Thomas shunt
CMR: cardiac magnetic resonance
CPS: critical pulmonary stenosis
EF: ejection fraction
LV: left ventricle
PA/IVS: pulmonary atresia with intact ventricular septum
PR: pulmonary regurgitation
PVR: pulmonary valve replacement
RV: right ventricle
RVEDV: right ventricular end-diastolic volume
TOF: tetralogy of Fallot
TR: tricuspid regurgitation

## Acknowledgements

None

